# Blockchain based clinical trial management system: A scoping review and conceptual framework

**DOI:** 10.1101/2024.06.12.24308813

**Authors:** M. C. Arjun, Ashikh Ahamed, Anura V. Kurpad, Tinku Thomas

## Abstract

**Objective:** The primary objective of the study was to map the real-world evidence of using Blockchain technology in clinical trial management systems and to create a novel conceptual framework for a Blockchain based digital ecosystem. As a first step towards validation of this conceptual framework, we aimed to create a blockchain simulation in Python programming language.

**Methods:** We did a scoping review of research articles which demonstrated a proof-of-concept or real-world application of Blockchain technology in clinical trial management. We searched in the PubMed, Web of Science, and IEEE Xplore to retrieve original articles published in English till April 2023. A novel conceptual framework was developed for a Blockchain based digital ecosystem with all the stakeholders involved in the conduct of clinical trial. We coded a simulation of clinical trial specific blockchain in Python using Django framework and the codes are made publicly available in GitHub.

**Results:** We retrieved 960 abstracts and included 21 full text articles in the review. Private blockchains like Hyperledger fabric and Ethereum are the most popular choice of platform. Smart contracts act as a key functionality in the Blockchain system to control specific activities. Clinical trial data was mostly stored outside the Blockchain, but Interplanetary File System is a popular choice for decentralized storage of data.

**Conclusions:** The scoping review, conceptual framework and the open-source Python codes would act as a guiding map for future research and product development in Blockchain based clinical trial management system and advancement in clinical research informatics.

## Introduction

Blockchain is an emerging technology which came into prominence after the introduction of Bitcoin in 2008.[1] Blockchain has already disrupted the financial sector and is finding new applications in various fields, including healthcare.[2–4] The key attraction of the blockchain is the distributed ledger system which means there is no requirement of a central authority to control the activities in the system.[5] This feature is augmented by Smart contracts, which are basically programming codes that can execute themselves when the programmed conditions are met.[6] The catchphrase - code is law - best summarizes the working of Smart contracts.[7] Although bulk data storage is still a challenge in blockchain systems, the key highlights of blockchain technology includes decentralization, better security, transparency, and auditability. The blockchain records are also immutable which ensures data integrity. In short, the blockchain system meets all the health informatics need of data provenance, privacy, integrity and is best suited for strict data management requirements that are needed in healthcare settings.[8]

The are multiple applications of blockchain proposed and tested in healthcare settings.[9] According to a survey conducted by the software company IBM, the greatest benefit of using blockchain will be in three areas: Clinical trials, regulatory compliance, and health records.[10] The focus of this study is on the application of blockchain in clinical trial management. Typically, a clinical trial is conducted to determine the efficacy of drugs and interventions and is mostly mandatory for approval of new drugs.

The entire process of conducting a clinical trial, starting from the protocol registration in trial registry to publication of results in peer reviewed journal, is based on trust in the investigators and the data they have generated. A meta-analysis of qualitative studies found that mistrust on trial organization as important barrier to clinical trial participation.[11] Similarly, regulatory bodies like the Food and Drug Administration (FDA) has identified the lack of traceability of clinical trial data as a major data management issue, reducing trust in the trial data. [12,13] The features of blockchain have the potential to validate this trust and provide a platform for tractable conduct of clinical trials which can be verified by any of the stakeholders, including trial participants. Since some of the technical concepts discussed here may not be familiar to the readers, we have given links for further reading in the supplementary material S1 Appendix.

The primary objective of this study was to conduct a scoping review to understand the current state of evidence on application of blockchain technology, including the smart contracts functionality, in the clinical trial management systems. Based on the learnings from this exercise, we discuss a novel conceptual framework for a digital ecosystem implementing blockchain for clinical trial management with different stakeholders that include regulatory bodies, ethics committee, and participants. As a first step towards validation of this framework, we tested a Blockchain simulation coded in Python programming language.

## Materials and Methods

### Methodology of the scoping review

We searched PubMed, Web of Science, and IEEE Xplore to retrieve titles and abstracts of peer-reviewed research papers. PubMed was searched systematically by combining key words and MeSH terms and the final search strategy was developed after multiple iterations. The search strategy for PubMed is given below. The articles published up to April 2023 were included in this review.

### PubMed search strategy

(“Blockchain”[MeSH Terms] OR “blockchain*”[Title/Abstract] OR “smart contract*”[Title/Abstract] OR “dapp”[Title/Abstract] OR “decentralized application*”[Title/Abstract] OR “peer to peer”[Title/Abstract] OR “cryptocurren*”[Title/Abstract] OR “Ethereum”[Title/Abstract] OR “Bitcoin”[Title/Abstract] OR “hyperledger”[Title/Abstract] OR “Proof of Work”[Title/Abstract] OR “Proof of Stake”[Title/Abstract] OR “Non-Fungible Token”[Title/Abstract] OR “Byzantine Fault Tolerance”[Title/Abstract]) **AND** (“Clinical Trial”[Publication Type] OR “Clinical Trials as Topic”[MeSH Terms] OR “trial*”[Title/Abstract] OR “clinical trial*”[Title/Abstract] OR “RCT”[Title/Abstract] OR “r c t”[Title/Abstract] OR “RCTs”[Title/Abstract])

The search results from all three databases were exported to the reference manager Zotero. Duplicates and retracted articles were removed at this stage. The references were further exported to the systematic review web-portal Rayyan.ai.[14] Rayyan duplicate removal tool was utilized to clear any additional duplicates and final set of references were ready for screening. Two reviewers (MCA and AA), who were blinded to each other’s decision, independently reviewed the titles and abstracts of all the articles based on the inclusion and exclusion criteria. We only included articles, including published conference proceedings, which demonstrated a proof of concept or real-world application of blockchain in clinical trials and which was published in English. Reviews, commentaries, or perspectives were excluded. In case of discrepancy between the reviewers, a senior author was consulted to decide on the inclusion of article into the review.

Once all disagreements among reviewers were sorted out, the final list of references with meta data was exported into a Microsoft Excel file for full text screening. The screening of full text was also based on the same inclusion exclusion criteria and was done blinded, and any discrepancy was resolved with help of senior author. Once the list of eligible articles was finalized, we proceeded to data coding and extracting which was done in Microsoft Excel file. The data extraction sheet was pre-tested before final use and is given in the supplementary material S2 Table.

### Development of Conceptual framework and testing in Python

To create a conceptual framework for blockchain based clinical trial management digital ecosystem, we first mapped all the stakeholders and systems involved in the conduct of a clinical trial. This was achieved using information available in the public domain, the literature review we conducted and talking to experts in clinical trial management. The next step was to conceptualize a new data workflow system using blockchain, which has the government regulatory bodies and systems at its core. The workflow is designed in such a way that private entities like clinical research organizations, data management solutions etc. have freedom to choose and operate their own blockchain ecosystems and at the same time ensure that data reporting and regulatory standards of the government are met.

As a first step towards validation of this framework, we tested the concept using a web portal designed to interface with a trial-specific blockchain. Developed on the Django Framework with Python, this prototype leverages encryption libraries, such as hashlib and cryptography, to ensure data integrity. The core infrastructure is an emulated file-based blockchain network, designed to synchronize the changes in one system to all other systems in the chain. This demonstrates important blockchain benefits, including immutability, full traceability, and transparency. We used the methodology of a clinical trial we did in our institute on Long COVID to emulate the working of a clinical trial. The case record form we used is given in the supplementary material S3 Table.

The detailed description of the methodology and Python codes are open sourced and posted in the GitHub repository (https://github.com/ictashik/BlockChain_ClinicalTrial). The description of the codes is also posted. Although the testing in Python demonstrates the feasibility of integrating blockchain principles into clinical trial processes through a web portal, this model is preliminary and not suited for production deployment.

## Results

### Results of Scoping review

We deployed the final search strategy in all the databases and retrieved a total of 960 references. After initial screening of titles and abstract, 57 articles were shortlisted, and full text was retrieved. After reading the full text articles and building consensus, we finalized 21 articles for inclusion in this scoping review. (Figure 1)

**Figure 1.**
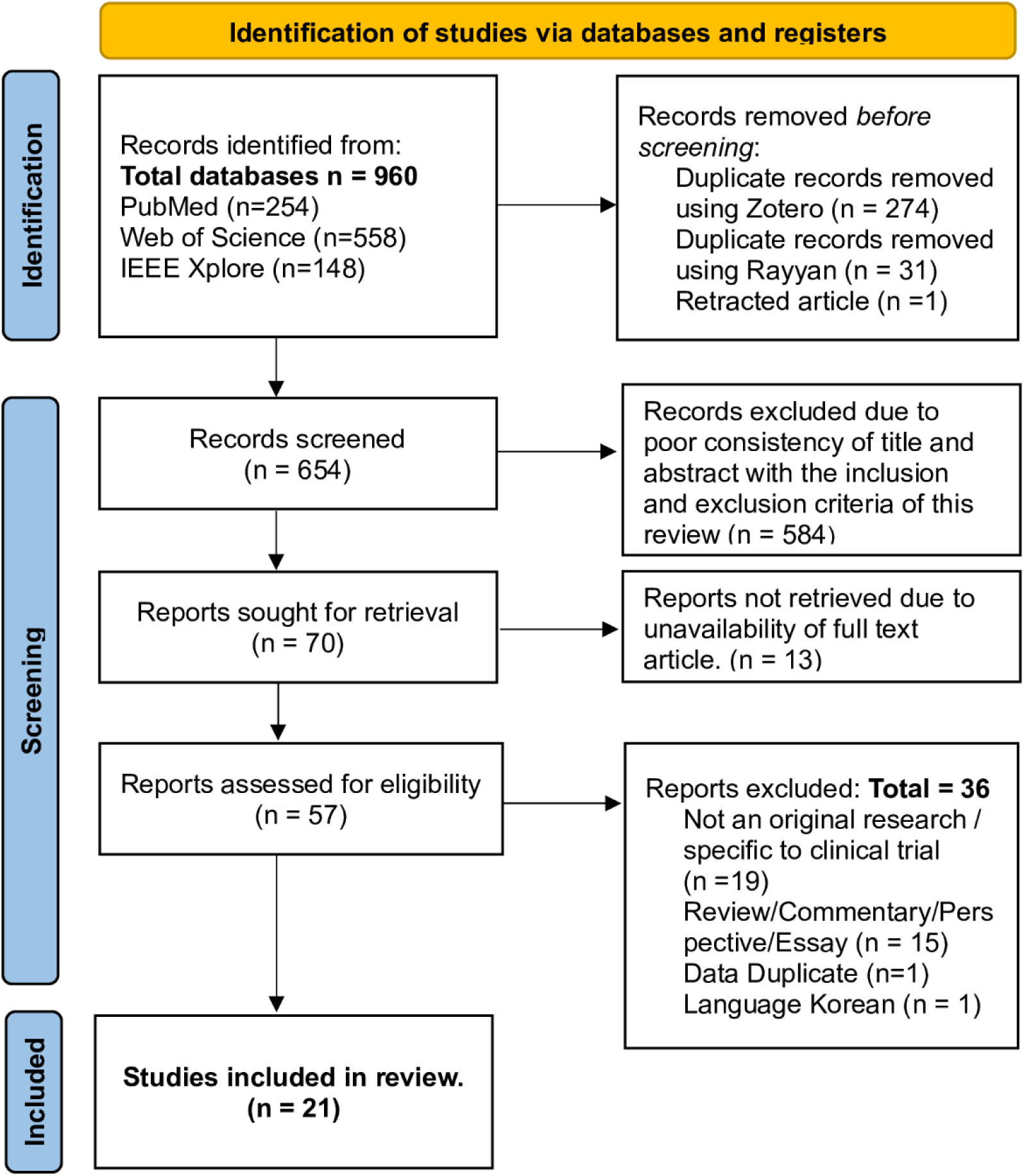
PRISMA flow diagram showing the scoping review process and selection of articles.

Table 1 shows the technical details of the blockchain systems used in their research. Hyperledger fabric and Ethereum, which are private blockchains, are the most used platforms. Private blockchain was preferred by almost all the researchers. Smart contracts are a key functionality present in almost all blockchain systems for controlling the activities in clinical trials like recruiting the participants, generating reports, and executing complex functions. Most of the blockchain based systems utilize off-chain storage of bulk trial data, or they have not specified. Interplanetary File System (IPFS) is one of the popular choices for decentralized storage of data. A short summary of the included articles is given in Table 2. Most of the research is proof-of-concept and deals with only a specific aspect part of clinical trial management like Dynamic consent or patient recruitment. Only one study tested the system, called METORY, in an actual clinical trial using virtual drugs and the same was registered in clinical trials registry.[15,16]

**Table 1.** Technical details of the blockchain in the included articles.

| **No .** | **First Author (Country; Year of Publication)** | **Blockchain: Name/Type/ Features** | **Smart contract** | **Trial data storage** |
| --- | --- | --- | --- | --- |
| 1 | Ki Young Huh (South Korea; 2022)[16] | Hyperledger Fabric (Private blockchain) | Chaincode (Smart contract) used for registering the consent and data transmission | External database – Amazon Relational Database Service |
| 2 | Yan Zhuang (China; 2022)[17] | Quorum blockchain, Private, Raft consensus mechanism | Multiple smart contracts used in the Clinical Trial Management system | Peer-to-peer distributed file system called the Interplanetary file system (IPFS) |
| 3 | Faisal Jamil (UK; 2022)[18] | Hyperledger Fabric v1.4.1 installed on Linux system | A smart contract is used to automate transactions. | Not specified / Not applicable |
| 4 | Lei Hang (China; 2021)[19] | Hyperledger Fabric (Permissioned blockchain) | Smart contract is written in JavaScript by using the Visual Studio Code | Off-chain data storage (Distributed data lake) |
| 5 | Baldwin C. Mak (Canada; 2021)[20] | Hyperledger Fabric v1.4 (private, permissioned) | Not specified | Not specified / Not applicable |
| 6 | Ivan da Silva Sendin (Brazil; 2021)[21] | VaccSC protocol implemented in Solidity language. Ethereum used | Smart contract is a core function of the application | Not specified / Not applicable |
| 7 | Yan Zhuang<br>(USA;<br>2020)[22] | Ethereum<br>(Private<br>blockchain) | Multiple smart<br>contracts used for<br>patient<br>recruitment,<br>engagement, and<br>monitoring | Clinical trial<br>database is<br>utilized.<br>Transaction<br>records are stored<br>in the blockchain. |
| 8 | Ilhaam A.<br>Omar<br>(United Arab<br>Emirates:<br>2020)[23] | Ethereum<br>(Private<br>blockchain) | Ethereum smart<br>contract using a<br>browser-based<br>compiler Remix<br>IDE | InterPlanetary File<br>System (IPFS) |
| 9 | Yan Zhuang<br>(USA;<br>2020)[24] | Ethereum<br>(Private<br>blockchain) | Smart contracts<br>are coded in the<br>layers of 3-layered<br>blockchain<br>architecture | Data was not<br>stored in<br>blockchain. Only<br>metadata is<br>created and<br>updated in the<br>blockchain. |
| 10 | Giuseppe<br>Albanese<br>(Switzerland;<br>2020)[25] | Hyperledger<br>Fabric<br>(Permissioned<br>blockchain) | Chaincode (smart<br>contract) used | Stored off-chain by<br>integration with<br>REDCap<br>database. |
| 11 | Tomonobu<br>Hirano<br>(Japan;<br>2020)[26] | Hyperledger<br>Fabric v1.0<br>(Linux<br>Foundation) | Chaincode (smart<br>contract) used. | Amazon Web<br>Services – off-chain data storage |
| 12 | Hans H. Jung<br>(Germany<br>2020)[27] | Ethereum based<br>Ocean<br>Blockchain | Service Execution<br>Agreements (SEA)<br>which are smart<br>contracts for<br>execution and<br>logging of<br>transactions. | Decentralized<br>database |
| 13 | Ilhaam A.<br>Omar (United<br>Arab Emirates;<br>2019)[28] | Ethereum<br>(Private<br>blockchain) | Written in Solidity<br>Language using<br>In-Browser<br>Remix IDE | InterPlanetary File<br>System (IPFS) |
| 14 | Olivia<br>Choudhury<br>(USA;<br>2019)[29] | Hyperledger<br>Fabric v1.4:<br>(Private<br>blockchain) | Multiple chaincode<br>(smart contracts)<br>for enrolment,<br>monitoring, Data<br>analysis/reporting. | Not specified |
| 15 | Yan Zhuang<br>(USA;<br>2019)[30] | Ethereum<br>(Private<br>blockchain) | A master smart<br>contract and<br>Multiple trial-based<br>smart contracts | Records will be<br>stored using trial<br>based smart<br>contracts. |
| 16 | Daniel R.<br>Wong (USA;<br>2019)[31] | Python and<br>Django software<br>development<br>framework | Not specified | Data storage and<br>control is outside<br>the blockchain |
| 17 | David M Maslove (Canada; 2018)[32] | Ethereum (Private blockchain) | Patient and Research smart contracts | Blockchain stores transaction records. Trial data is stored off-chain. Interaction is managed by a blockchain "oracle". |
| 18 | Yan Zhuang (USA; 2018)[33] | Private blockchain simulation | Multiple smart contracts coded in the private blockchain simulation | Remote Procedure Call (RPC) server is used as a bridge to connect clinical sites databases to perform information exchange |
| 19 | Mehdi Benchoufi (France; 2017)[34] | Bitcoin network, Consensus by Proof-of-Process concept | Not used | Chainscript document stored all the data |
| 20 | Fabio Angeletti (Italy; 2017)[35] | Ethereum (Private blockchain) | Not used in Proof of concept | Not specified |
| 21 | Timothy Nugent (UK; 2016)[36] | Ethereum (Private blockchain) | Hierarchical arrangement of two core types of smart contract written in JavaScript and Solidity | InterPlanetary File System (IPFS) or Ethereum's native Swarm protocol |

**Table 2.** Short summary of the included articles in the scoping review.

| No. | First Author (Country; Year of Publication) | Short summary |
| --- | --- | --- |
| 1 | Ki Young Huh (South Korea; 2022)[16] | The paper describes the development of a Dynamic consent platform for clinical trials called METORY. The system was tested in an actual clinical trial using virtual drugs. (ClinicalTrials.gov identifier: NCT05047016) |
| 2 | Yan Zhuang (China; 2022)[17] | The main objective was to reengineer the traditional Clinical Trial Management system (CTMS) using Blockchain that spans all stages of clinical trial. Case studies were conducted to assess feasibility, scalability, stability, and efficiency of the proposed blockchain based CTMS. |
| 3 | Faisal Jamil<br>(UK; 2022)[18] | This paper proposes a real-time network feedback model and uses a clinical trial service framework to test the network and evaluate the effectiveness of the designed model. |
| 4 | Lei Hang<br>(China; 2021)[19] | This paper proposed a blockchain platform with smart contracts for clinical trial management that ensures traceability, automation, and prevention of posteriori reconstruction. A proof of concept is implemented and efficiency, and usability is demonstrated. |
| 5 | Baldwin C. Mak<br>(Canada; 2021)[20] | This study conducted a proof-of-concept pilot study using blockchain in an ongoing clinical trial. This parallel group study aimed to assess the impact on blockchain on multiple end points including visit time, cost, compliance, and user experience when compared with the standard methodology. |
| 6 | Ivan da Silva Sendin<br>(Brazil; 2021)[21] | This paper proposed a smart contract enabled blockchain system, named VaccSC, which can improve the conduct and auditability of Phase III vaccine clinical trials. |
| 7 | Yan Zhuang<br>(USA; 2020)[22] | The paper describes a comprehensive blockchain framework for Virtual Clinical Trial, which is a new and innovative method for home based clinical trials. The simulations and case studies demonstrate the working of the system on patient recruitment, engagement, and monitoring. |
| 8 | Ilhaam A. Omar<br>(United Arab Emirates; 2020)[23] | The objective of the research is to provide a working proof-of-concept for application of blockchain technology in clinical trials. The research concludes that the proposed framework is highly effective in ensuring data integrity, transparency, and traceability. |
| 9 | Yan Zhuang<br>(USA; 2020)[24] | The primary objective of this research was to create a generalized blockchain architecture that provides data coordination functions which can be used for a wide range of health care applications. A clinical trial recruitment scenario was simulated to test the feasibility of the application. |
| 10 | Giuseppe Albanese<br>(Switzerland; 2020)[25] | The paper presents an approach called SCoDES, which is made available as a self-contained infrastructure, that utilize blockchain technology for managing dynamic consent in clinical trials. |
| 11 | Tomonobu Hirano<br>(Japan; 2020)[26] | The main objective of the paper was to validate a system that uses blockchain to enable data security in clinical trials. The system was developed, validated, and tested for its resilience to data tampering. |
| 12 | Hans H. Jung<br>(Germany 2020)[27] | This paper discusses a Decentralized Clinical Study Consent Management which creates, manages, and store consent documents in a decentralized way. All changes are recorded on-chain and logs can be viewed and verified. |
| 13 | Ilhaam A. Omar (United Arab Emirates; 2019)[28] | A proof-of-concept study designed to use blockchain for clinical trial data management. The framework captures three different stages of clinical trial, and interaction among multiple stakeholders. |
| 14 | Olivia Choudhury (USA; 2019)[29] | A blockchain based data management system is designed and implemented for multi centric clinical trials. A comparative performance analysis and comprehensive study is conducted to demonstrate the value and effectiveness of the proposed system. |
| 15 | Yan Zhuang (USA; 2019)[30] | The paper proposes a blockchain model containing multiple smart contracts for clinical trial management and patient engagement. The focus of the research is on patient recruitment. |
| 16 | Daniel R. Wong (USA; 2019)[31] | A proof-of-concept web portal was created, and real data was used to simulate the progress of a clinical trial. The system's capability for providing a trace for data flow and audibility was assessed. The system demonstrates better management of clinical trial data and an improvement in the adverse event reporting. |
| 17 | David M Maslove (Canada; 2018)[32] | The study developed a proof-of-concept system, called BlockTrial, for data management in clinical trials. The system promote transparency and increase trustworthiness of data, benefiting all stakeholders, including patients. |
| 18 | Yan Zhuang (USA; 2018)[33] | This paper investigates the workflow of Health Information exchange and clinical trials and uses a private blockchain model to tackle some of the issues. The work is a proof of concept providing simulations of potential solutions using blockchain and smart contracts to monitor clinical trials. |
| 19 | Mehdi Benchoufi (France; 2017)[34] | A proof-of-concept protocol was designed to record the consent of participants in blockchain. The consent was sought at every point of protocol change. The time stamped consent process was obtained in single document which can even be verified by the public. |
| 20 | Fabio Angeletti (Italy; 2017)[35] | A proof of concept and its implementation, and performance analysis for a digital system which combines Internet of Things and Blockchain. The focus of is on clinical trial recruitment with privacy preserving flow of personal data. |
| 21 | Timothy Nugent (UK; 2016)[36] | This paper introduces a system of blockchain which uses smart contracts to improve trust and address the issues of data manipulation in clinical trials. The blockchain and smart contract will act as a trusted administrator keeping an immutable record of trial history and data. |

### Conceptual framework and testing in Python

Figure 2 shows the newly developed conceptual framework for a Clinical Trial Management Digital Ecosystem. At the center of the proposed framework is the clinical trial registry. We envision a system where all stakeholders interact with the central registry through a blockchain using smart contracts. Fundamentally there should be three systems which have separate login credentials through the registry frontend. A view only version for the public will also be available like the present registry.

**Figure 2.**
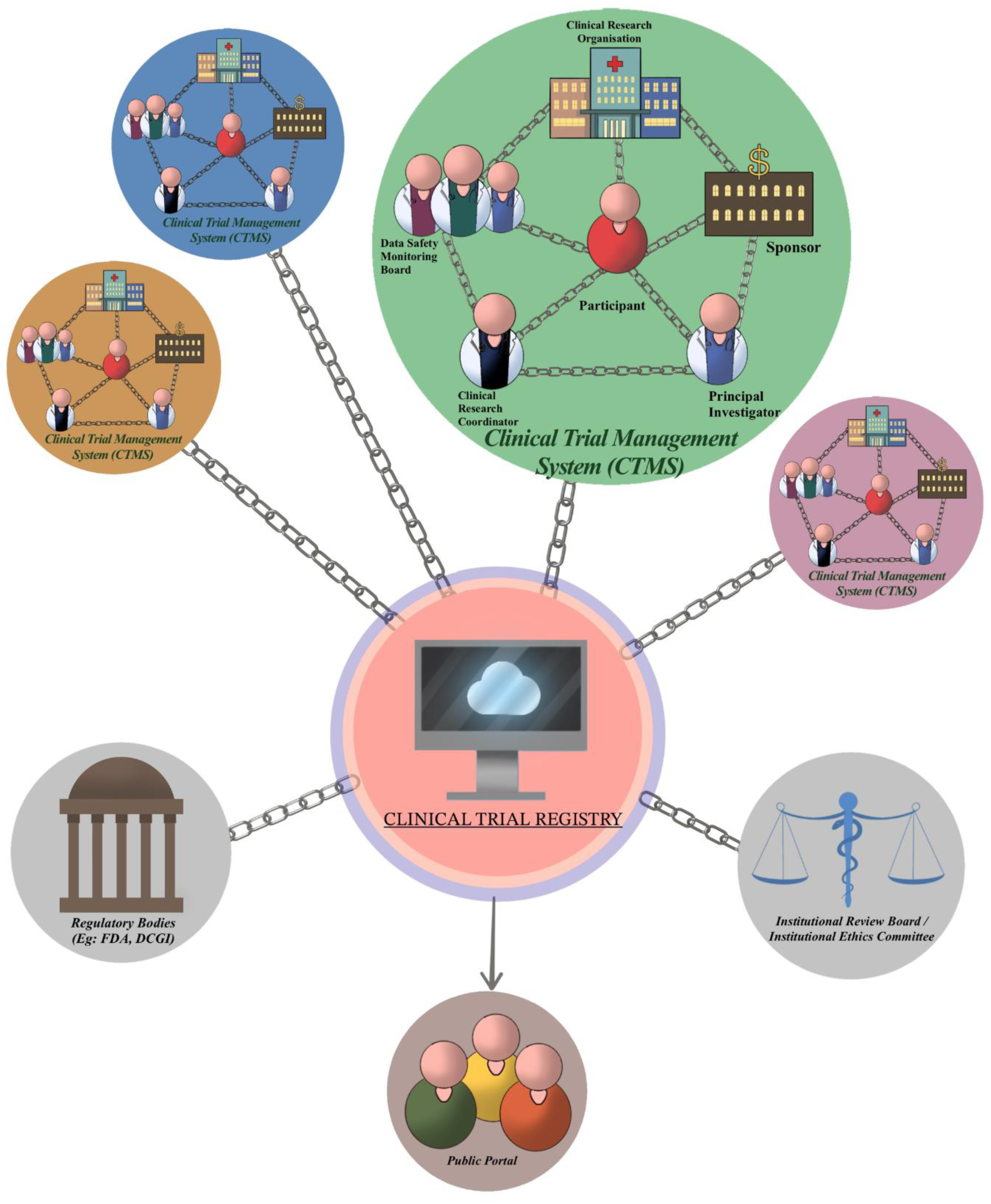
Conceptual framework for Clinical Trial Management Digital Ecosystem.

The first type of system in the figure is represented by blockchain based trial management system with stakeholders who are directly involved in the conduct of the trial: Sponsor, Clinical Research Organization, Principal Investigator, Clinical Research Coordinator, Data Safety Monitoring Board, and the trial participants. This system can be standalone and managed by private entities providing blockchain based trial management services. All trial related data will be registered on the blockchain. The other entity in the framework includes institutional review boards or ethics committee and Regulatory bodies (E.g.- Food and Drug Administration). All the entities are connected and can interact with each other through the central blockchain based database of clinical trial registry.

The interactions, data exchange, and interoperability will be managed by smart contracts. For example, before the trial begins, the protocol in the clinical trial management system should be synced with the registry and approval of ethics committee and regulatory bodies must be obtained. The approvals can be given using digital signatures and this will be timestamped in blockchain. Any change in the protocol will be automatically reflected in the registry for approval and further action. The progress of the trial will be automatically synced to the registry and a public view portal will display the progress summary of the trial automatically. (E.g., The number of participants enrolled) If the regulatory bodies need access to raw data for further auditing purposes, they can query the clinical trial management system through central registry blockchain in compliance with the smart contracts.

As a first step towards validation of this framework, we created a simulation of Blockchain based clinical trial system and the codes for the same can be found in https://github.com/ictashik/BlockChain_ClinicalTrial. The code is open sourced to encourage reproducibility.

## Discussion

We did a scoping review to identify the current developments in the blockchain based clinical trial management systems. Most of the studies have focused on reimagining the traditional clinical trial management system, in part or whole, by replacing the centralized management of data with the decentralized blockchain and smart contracts. While most of the studies were at a pilot stage or proof-of-concept, one research group has launched a clinical trial exclusively to study their system using virtual drugs. [15,16, 37] Although not addressed by majority of studies, the challenges of implementing a blockchain system for clinical trials are well identified in the literature.[38,39]

The scoping review found that Private or Permissioned blockchain platforms like Hyperledger fabric and Ethereum are most popular choices for software development. This is consistent with its overall popularity in healthcare based blockchain systems.[40] Although it can be argued that private or permissioned blockchain reduces the claim of decentralization, this is ideal for the data management issues like privacy, and data security present in clinical trials.[31,41] Similarly smart contracts are central to all systems and the process of decentralization is incomplete without smart contracts.[42] Data management in blockchain has its own benefits and challenges, especially when dealing with big data.[43] The InterPlanetary File System (IPFS) is considered as a promising solution to decentralised data storage.[44] The IPFS system was used in few of the articles included in the review. [17,23,28,36]. There are cost implications also in developing a blockchain based system, which are not discussed or accounted in most research papers.[45]

To the best of our knowledge, the conceptual framework we developed is unique because it takes a holistic approach by involving the government systems and regulatory bodies in the blockchain based clinical trial management digital ecosystem. A research organization conducting a clinical trial is given freedom to choose their own private software solution but ultimately the data is converged in the central clinical trial registry for approvals and audit. The blockchain technology ensures that all the data management issues are addressed which includes data provenance, transparency, and auditability. This framework will help to reimagine and revolutionize the conduct of clinical trial and guide government organizations working to bring blockchain in public domain.[46]

As a first step towards validation of the conceptual framework, we tested the idea in a simulation in Python and the codes are made public for reproducibility. In real world setting, the interoperability of the blockchain systems or decentralized apps (DApps) can be ensured using networks which allow cross-chain transfer of data.[47] The data standards currently used in healthcare setting can be adopted for this purpose as well. Along with the scoping review which maps the evidence in the literature, the conceptual framework and the simulations are a major strength and contribution of this paper.

This study had some limitations. We could not retrieve the full text of all the articles we marked eligible during the screening process, and this could bias our results. The conceptual framework did not consider the cost implications of developing and maintaining such a system. We have not explored the convergence of the rapidly evolving field of artificial intelligence into our blockchain framework.

## Conclusions

The internet is changing, and the blockchain technology is widely regarded as the future of internet. These changes provide tremendous opportunity to improve or even revolutionize the conduct of clinical trial and the progress of science as well. We hope this paper will inspire ideas and provide a roadmap for blockchain based systems and product development in the field of clinical trials.

## Acknowledgements

We thank Mr. G.G. Romould for Figure 2 artwork.

## Data Availability

All data related to this study are available in the article or as supplementary material. Python codes of Blockchain simulation are available at: https://github.com/ictashik/BlockChain_ClinicalTrial

## CRediT Authorship Contribution Statement

**MCA**: Conceptualization, Methodology, Validation, Formal analysis, Investigation, Data Curation, Visualization, Project administration, Writing - Original Draft, Writing - Review & Editing. **AA**: Software, Methodology, Validation, Formal analysis, Investigation, Data Curation, Visualization, Writing - Review & Editing. **AVK**: Validation, Methodology, Formal analysis, Visualization, Resources, Supervision, Funding acquisition, Writing - Review & Editing. **TT**: Validation, Methodology, Formal analysis, Resources, Data Curation, Supervision, Funding acquisition, Writing - Review & Editing

## upplementary Material

**S1 Appendix.** Wikipedia links for further reading

**S2 Table.** List of variables in the data extraction sheet and their description

**S3 Table.** Clinical trial case record form used for Blockchain simulation

