## supplementary material for "Blockchain based clinical trial management system: A scoping review and conceptual framework"

#### S1 Appendix - Wikipedia links for further reading

- Blockchain: https://en.wikipedia.org/wiki/Blockchain
- Clinical trial management system: https://en.wikipedia.org/wiki/Clinical_trial_management_system
- Smart contract: https://en.wikipedia.org/wiki/Smart_contract
- InterPlanetary File System: https://en.wikipedia.org/wiki/InterPlanetary_File_System
- Bitcoin: https://en.wikipedia.org/wiki/Bitcoin
- Ethereum: https://en.wikipedia.org/wiki/Ethereum
- Hyperledger: https://en.wikipedia.org/wiki/Hyperledger
- Distributed ledger: https://en.wikipedia.org/wiki/Distributed_ledger

### S2 Table

**S2 Table. List of variables in the data extraction sheet and their description**

| SN | Data item | Description |
|---|---|---|
| 1 | Year | Year of publication of the article |
| 2 | First author | First author of the article |
| 3 | Country | Country of the first author |
| 4 | Study objective | Objective of the research with a real-world application |
| 5 | Study design | Proof of concept/Clinical trial/application |
| 6 | Name of blockchain | Hyperledger/Ethereum/Bitcoin/Other |
| 7 | Type of blockchain | Private/Consortium/Public |
| 8 | Consensus mechanism | Proof of Work/Proof of stake/Other |
| 9 | Use of smart contract | Yes/No - with description |
| 10 | Data storage | Centralized or decentralized data storage |
| 11 | Summary / Conclusion | The summary or conclusion of the proposed solution |
| 12 | Research contribution | Our appraisal of the proposed solution |

### S3 Table

**S3 Table. Clinical trial case record form used for Blockchain simulation***

| Q.No. | Question | Answer |
|---|---|---|
| | **ENROLEMENT** | |
| 1 | Do you have the Long COVID symptom of fatigue? | Yes / No (End form) |
| 2 | Consent | Yes / No (End form) |
| | **BASELINE Data** | |
| 3 | Participant enrolment number | LC001 |
| 4 | Group (Allotted by randomization) | A / B |
| 5 | Date of Enrolment | DD/MM/YYY |
| 6 | Age in completed years | Integer |
| 7 | Sex | Male/Female/Other |
| 8 | Highest level of education completed? | Open |
| 9 | Do you have history of allergy? | Yes / No |
| 10 | Did you receive Covid-19 vaccine? | Yes / No |
| 11 | Any diagnosed disease or co-morbidities? | Yes / No |
| | **FOLLOW-UP after 1 month** | |
| 12 | Date of final follow-up | DD/MM/YYY |
| 13 | Number of antihistamine tablets taken: | Integer |
| 14 | Do you have the Long COVID symptom of fatigue during the final follow-up? | Yes / No |

*Title of the study used for simulation**:** Antihistamines for treatment of Long COVID fatigue – A placebo controlled double blinded Randomized Controlled Trial

Simulation link: https://github.com/ictashik/BlockChain_ClinicalTrial
